# Factors Influencing the Success of Community-Based Participatory Approach in Implementing an HIV Stigma Reduction Intervention in Indonesia

**DOI:** 10.64898/2025.12.23.25342924

**Authors:** Mona H Sheikh Mahmud, Rafdzah A Zaki, Theresia P. Kusumoputri, Devika Devika, Dean Retno, Frederick L Altice, Adeeba Kamarulzaman

**Author notes:** Corresponding author: Rafdzah A Zaki. These authors contributed equally to this work (Mona H Sheikh Mahmud, Rafdzah A Zaki, Adeeba Kamarulzaman). These authors have contributed equally to this work (Theresia P. Kusumoputri, Devika Devika, Dean Retno, Frederick L Altice).

## Abstract

**Background:** HIV-related stigma among healthcare providers hinders service delivery and patient engagement, especially in low- and middle-income countries. The *Intervensi Penghapusan Stigma dan Diskriminasi* (IPSD) intervention employs a Community-Based Participatory Approach (CBPA) to reduce stigma among healthcare workers (HCWs) in Indonesia by involving people with HIV (PWH) and key populations (KP) as co-developers and co-implementers.

**Methods:** This cross-sectional study evaluated the implementation outcomes of adoption and fidelity, as defined by Proctor et al., using a validated 5-point Likert scale survey developed based on the Consolidated Framework for Implementation Research (CFIR) and Proctor et al. framework. A total of 120 physicians, nurses, midwives, and laboratory technicians from 31 primary health centres (PHCs) in Greater Jakarta participated in the survey. PHCs were categorised as either high- or average-performing based on triangulated data from PWH networks and evaluations by the Ministry of Health. Descriptive statistics and bivariate analyses, including chi-square and t-tests, were conducted at a significance level of p < 0.05.

**Results:** Significant association was found between occupational role and PHC performance (*p*=0.012). High-performing PHCs reported stronger technical expertise (*p*=0.033) and better HIV/STI epidemiological knowledge (*p*=0.033). Organisational incentives influenced fidelity (*p*=0.032), with higher-performing PHCs reporting greater institutional support.

**Conclusion:** Findings underscore the need to reduce stigma through equitable services and supportive organisational climates. Agreement across PHCs showed shared recognition of involving PWH and KP as co-implementers and facilitated intervention adoption, aligning with evidence for contact-based stigma reduction. Differences between PHCs were shaped by capacity and knowledge, with higher-performing facilities showing stronger intervention fidelity. Organisational incentives facilitated sustained fidelity, while national mandates ensured adoption. By examining CFIR constructs and Proctor outcomes, this study informs scalable stigma reduction in primary healthcare. Limitations include small sample size, limited scope, self-reported data, and cross-sectional design, precluding multivariable modelling, making findings exploratory.

## Introduction

In Indonesia, an estimated 570,000 people with HIV (PWH) in 2022, with a prevalence of approximately 0.4%(1). The most recent cascade data show that 60% of PWH know their status, 41% are receiving antiretroviral therapy (ART), and only 38% have achieved viral suppression, reflecting variations in data sources and measurement definitions(1). Although new HIV infections declined between 2021 and 2024, AIDS-related mortality increased, largely due to gaps in treatment coverage and in viral load monitoring. In response, the national HIV program prioritised the decentralisation of services from tertiary hospitals to primary health centres (PHCs) to improve testing, treatment initiation, and retention in community settings(2).

Health-related stigma by clinicians refers to their labelling, stereotyping, separating, status loss, and discrimination operating within power imbalances, which restrict access to prevention and care, increasing mortality and morbidity(3, 4). Even when a condition has limited clinical severity or is well controlled, stigma can still cause substantial harm, including psychological distress, social isolation, delayed care seeking, reduced treatment adherence, and diminished participation in education and work, which together lower the overall quality of life(5, 6).

A 2023 assessment of stigma among PWH in Indonesia found that 13.4% of PWH reported experiencing stigma and discrimination in the past year, with stigma from healthcare workers being common in HIV-specific (19.5%) and general healthcare services (15.9%)(7). Key populations (KP), including people who use drugs (16.8%), transgender individuals (16.7%), and female sex workers (12.6%), reported avoiding healthcare facilities due to fear of discrimination(7). Additionally, 58.9% of KP respondents delayed ART initiation due to concerns about their HIV status(7).

In 2018, Indonesia’s Ministry of Health introduced the evidence-based IPSD intervention to promote stigma-free HIV services, utilizing the FHI360 USAID-funded "Linkages Health4All: Training Health Workers for the Provision of Quality, Stigma-Free HIV Services for Key Populations" program(8). Since 2016, Linkages has conducted Health4All training for over 800 participants across 14 countries, continuously refining the curriculum based on field insights(9). Pre- and post-assessments consistently demonstrated enhanced knowledge and more positive attitudes toward key populations. Participants reported a better understanding of key populations, their vulnerabilities, and the challenges faced by both communities and providers(9). Governments in Cambodia, Indonesia, Kenya, and Suriname have since adapted and endorsed Health4All as a national training curriculum(9). This guide was tailored to the Indonesian context, locally named *"Intervensi Penghapusan Stigma dan Diskriminas"* (IPSD), and delivered through in-person training at 42 primary health centres (PHCs) and online training at 56 referral hospitals in Greater Jakarta, reaching 1,105 PHC staff and 2,057 hospital staff.

The IPSD intervention employed a Community-Based Participatory Approach (CBPA), engaging PWH and KP experts in adapting training modules and delivering training to HCWs. CBPA, recognised as the “gold standard” in health promotion and disease prevention among socio-economically deprived populations for improving health behaviours, service access, and literacy(10), has also been shown to be cost-effective in multi-stakeholder stigma reduction interventions involving PWH and key populations across seven South Asian countries(11), integrating community systems across all levels of healthcare from policy to service delivery(12, 13).

The role of the CBPA strategy in supporting IPSD has yet to be evaluated, offering an opportunity to assess its impact on implementation and potential for expansion. This study explored the factors contributing to the success of CBPA in facilitating the adoption and implementation fidelity of the IPSD intervention. It examines organisational characteristics, external influences such as policies and incentives, and key process indicators. Guided by Proctor et al.’s Implementation Outcomes Framework(14) and CFIR constructs(15), this study identifies barriers and facilitators critical to effective implementation.

## Materials and Methods

### Study Setting

This cross-sectional study used a 43-item validated survey based on CFIR domains and Proctor’s Adoption and Fidelity outcomes, administered to physicians, nurses, midwives, and laboratory technicians across 42 PHCs in Greater Jakarta who had completed IPSD training. The instrument, developed in Phase 2 of a doctoral study on IPSD implementation in Indonesia(16), underwent face, content, and cross-cultural validation through expert review and stakeholder feedback. Reliability analysis showed internal consistency (Cronbach’s alpha: α=0.637–0.945, ITC: 0.237–0.945, ICC: 0.440–0.967).

### Sample Size Calculation and Sampling Strategy

Using OpenEpi, a proportionate sample size of n=286 was calculated from the 1,105 trained personnel, assuming 50% adoption with fidelity and a 95% confidence level. Stratified random sampling was applied to ensure subgroup representation by age, gender, occupational role, and workplace, with random selection within each stratum.

### Selection of High and Average-Performing PHCs

As the IPSD intervention had not been previously evaluated, a new process was developed to classify PHCs based on their performance. High-performing (HP) and average-performing (AP) PHCs were identified through triangulated data from three sources: 1) the 2022 Jaringan Indonesia Positif (JIP) Survey assessing PWH and KP experiences; 2) the 2020 Indonesian AIDS Coalition (IAC) Survey on service accessibility; and 3) Ministry of Health (MoH) evaluations. HP- and AP- PHCs were classified using triangulated data from PWH and KP experience surveys(17, 18) and the MoH performance assessment(16). Facilities scoring ≥3 on the JIP 5-point scale or rated as “community-friendly” by the IAC and selected by the MoH were categorised as HP-PHCs, ensuring a balanced representation of both community and institutional perspectives (see Appendix 1).

### Measures and Analysis

The survey used a 5-point Likert scale (ranging from *strongly agree* to *strongly disagree*) was completed by physicians, nurses, midwives, and laboratory technicians from 31 PHCs in Greater Jakarta providing HIV services (see Appendix 2). The survey items covered three CFIR domains: Inner Setting, Outer Setting, and Process. It encompassed five constructs (Implementation Climate, Patient Needs and Resources, External Policy and Incentives, Engaging, Executing) and five sub-constructs (Tension for Change, Compatibility, Organisational Incentives and Rewards, External Change Agents). Of the total items, 22 were designed to measure adoption, while 21 were intended to assess fidelity (Figure 1).

**Fig 1.**
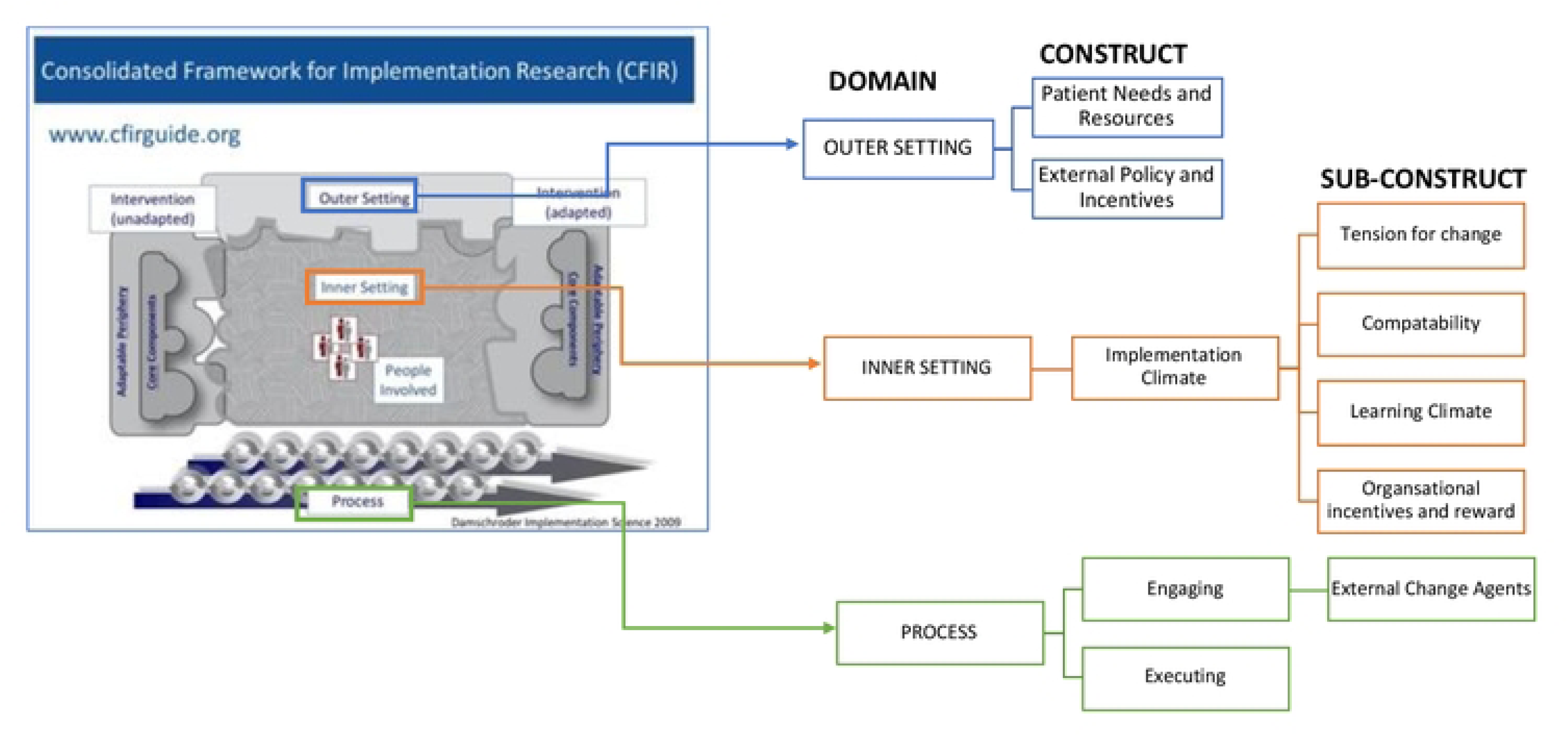
Selected CFIR Domains, Constructs and Sub-Constructs(19)

Data were collected via the KoboCollect online survey platform from July 3 to August 1, 2024, with approval from the MoH Indonesia and disseminated by the Heads of PHCs. Respondents received an equivalent of US$4 internet package as a token of appreciation for their participation. Data analysis involved frequency distributions, means, standard deviations, and bivariate analyses, with significance assessed using Chi-square and independent sample t-tests. All statistical tests were conducted using SPSS Version 29, with significance set at *p* < 0.05.

### Ethics Approval

This study was reviewed and approved by the Ethics Committee of the Institute of Research and Community Service, Atma Jaya Catholic University, Indonesia. All participants were informed of the purpose, procedures, and voluntary nature of the study. Written informed consent was obtained from all the participants. Confidentiality and anonymity were ensured throughout the research process, in accordance with the ethical guidelines for human subjects’ research.

## Results

### Respondents Characteristics

Although the study sought to recruit 286 HCWs from 42 PHCs, only 242 (80.4%) participants from 31 PHCs responded, with only 120 meeting the inclusion criteria and completing the survey. Although repeated recruitment efforts were undertaken, the target sample size was not achieved. Eligible participants included physicians, nurses, midwives, and laboratory personnel who had received IPSD training.

Most respondents were female (n=91, 75.8%), with representation across occupational roles: nurses (n=47, 39.2%), doctors (n=34, 28.3%), laboratory technicians (n=23, 19.2%), and midwives (n=16, 13.3%). Participants were from districts in East (n=41, 34.2%), South (n=44, 36.7%), North (n= 7, 5.8%), West (n=16, 13.3%), and Central Jakarta (n=12, 10.0%), with 46.7% (n=56) from HP-PHCs and 53.3% (n=64) from AP-PHCs. Facility performance was evaluated based on patient satisfaction, service quality, and institutional assessment by the MoH(16).

The participants’ occupational roles were significantly correlated with the performance of PHCs (*p*=0.012). Specifically, a higher proportion of nurses and doctors were observed in high-performing PHCs, whereas midwives and laboratory technicians were more prevalent in average-performing PHCs. Although geographic variations were observed, the differences across districts were not statistically significant (Table 1).

**Table 1:** Association between Respondents’ Characteristics and Performance of PHCs (N=120)

| Items | Variable | Performance |  | *p*-value |
| --- | --- | --- | --- | --- |
|  |  | High n (%) | Average n (%) |  |
| **Age category** | 25-35 | 39 (52) | 36 (48) | 0.066 |
|  | 36-45 | 13 (40.6) | 19 (59.4) |  |
|  | 46-55 | 2 (18.2) | 9 (81.8) |  |
|  | 56-65 | 2 (100) | 0 (0.00) |  |
| **Gender** | Female | 38 (41.8) | 53 (58.2) | 0.120 |
|  | Male | 17 (60.7) | 11 (39.3) |  |
|  | Transgender | 1 (100) | 0 (0) |  |
| **Job Title** | Nurse | 28 (59.6) | 19 (40.4) | 0.012 |
|  | Doctor | 18 (52.9) | 16 (47.1) |  |
|  | Midwife | 5 (31.3) | 11 (68.8) |  |
|  | Laboratory Technician | 5 (21.7) | 18 (78.3) |  |
| **District** | North Jakarta | 5 (71.4) | 2 (28.6) | 0.123 |
|  | East Jakarta | 23 (56.1) | 18 (43.9) |  |
|  | South Jakarta | 16 (36.4) | 28 (63.6) |  |
|  | West Jakarta | 5 (31.3) | 11 (68.8) |  |
|  | Central Jakarta | 7 (58.3) | 5 (41.7) |  |

### Adoption Outcomes

#### Frequency Distribution Reported Against CFIR Constructs and Sub-Constructs

Overall, the frequency distribution revealed strong support for the IPSD intervention across all CFIR domains, underscoring its perceived relevance, compatibility, and value in reducing stigma within primary healthcare settings (see Appendix 3).

#### Tension for Change

Most respondents viewed IPSD as necessary to strengthen linkages between PHCs and PWH support groups (n=115, 95.8%) and to address existing knowledge gaps in HIV, STIs, and KP needs (n=111, 92.5%), which were seen as underlying drivers of discriminatory behaviours (n=110, 91.7%). These findings suggest a collective awareness among HCWs of the need for IPSD training to enhance the quality of care and foster more equitable provider–patient relationships.

#### Compatibility

Most respondents perceived IPSD as being well aligned with PHC culture (n=106, 88.3%) and workflows (n=100, 83.3%), with minimal conflict with personal values (n=48, 27.0%). Its distinctiveness from previous training (n=111, 92.5%) and relevance to daily practice (n=105, 87.5%) suggest that IPSD is both acceptable and adaptable to existing service delivery structures.

#### Organisational Incentives and Rewards

Incentivised support for participation appeared uneven, with just over half (n=71, 59.2%) reporting incentives. This variation suggests potential organisational barriers that could influence consistent implementation and long-term sustainability.

#### Patient Needs and Resources

Stigma reduction emerged as a central institutional priority (n=108, 90%), with IPSD viewed as effective for improving patient care (n=117, 97.5%), reducing stigma (n=113, 94.2%), and supporting treatment uptake and retention (n=110, 91.7%). These results reflect the IPSD’s perceived success in addressing critical service delivery gaps for PWH and KPs.

#### External Policy and Incentives

Government mandates were identified as a key driver of IPSD adoption (n=104, 86.7%), demonstrating strong alignment between national policy frameworks and local implementation.

#### External Change Agents

External Change Agents are individuals or entities outside an organisation that influence innovation adoption(19). In this study, these agents included PWH and KP community experts who co-developed and co-implemented IPSD. PWH and KP representatives’ involvement was highly valued in development (n=108, 90%) and implementation (n=104, 86.7%), serving as facilitators who enhanced understanding (n=103, 85.9%) and reinforced skills (n=111, 92.5%). This community involvement demonstrates CBPA’s effectiveness in fostering partnership between healthcare workers and affected communities.

#### Association between Survey Items Measuring Adoption and PHCs Performance

No significant associations were found between PHC performance and CFIR constructs for adoption, suggesting comparable perceptions across facility types. Respondents from HP- and AP-PHCs recognised the intervention’s importance in strengthening PHC and community network linkages, aligning with organisational culture, and improving service quality. While incentive perceptions varied, most viewed stigma reduction and patient-centred care as priorities. Government mandates and PWH/KP trainer involvement were considered critical adoption facilitators. These findings indicate strong institutional readiness to adopt IPSD across PHCs, regardless of their performance (Table 2).

**Table 2:**
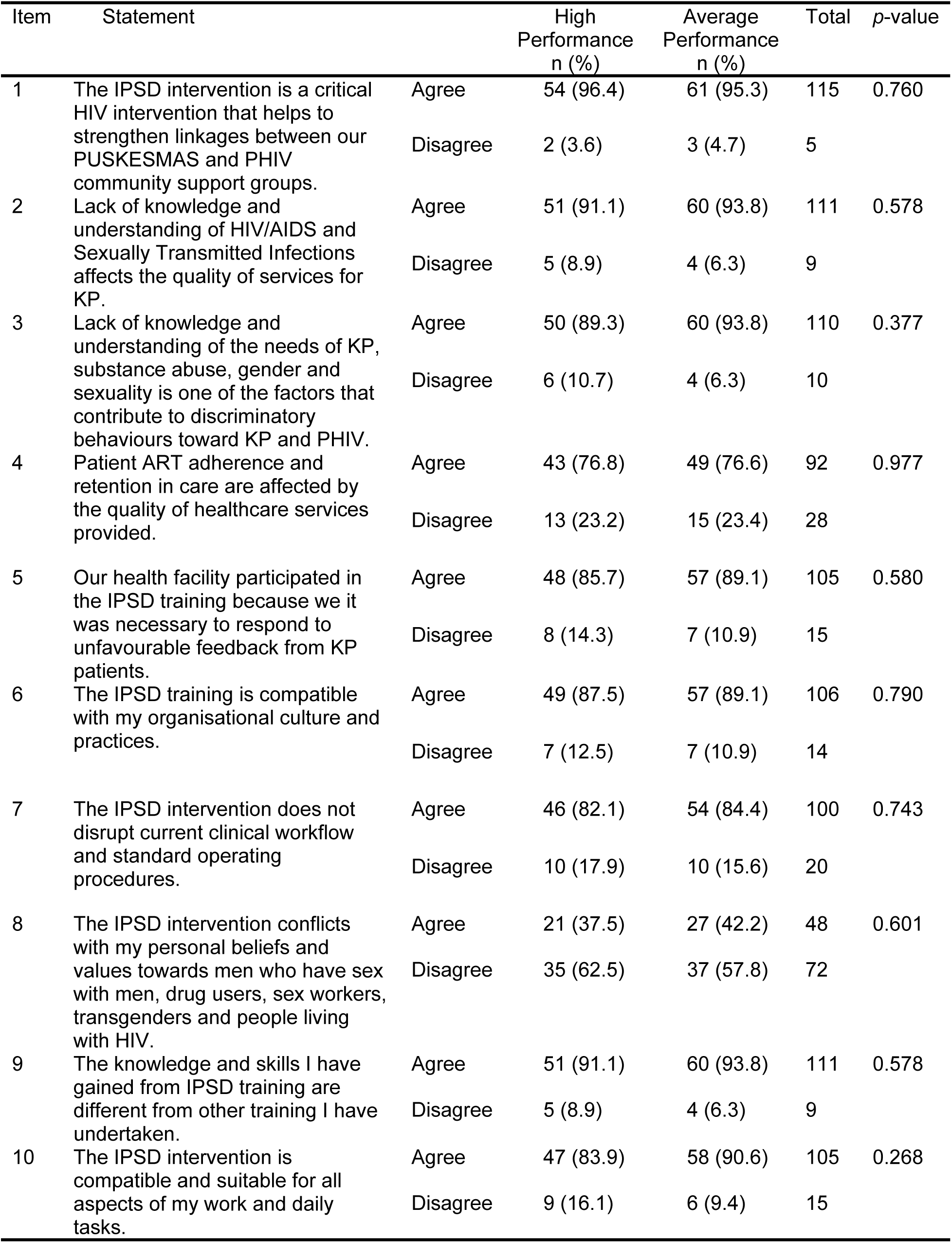
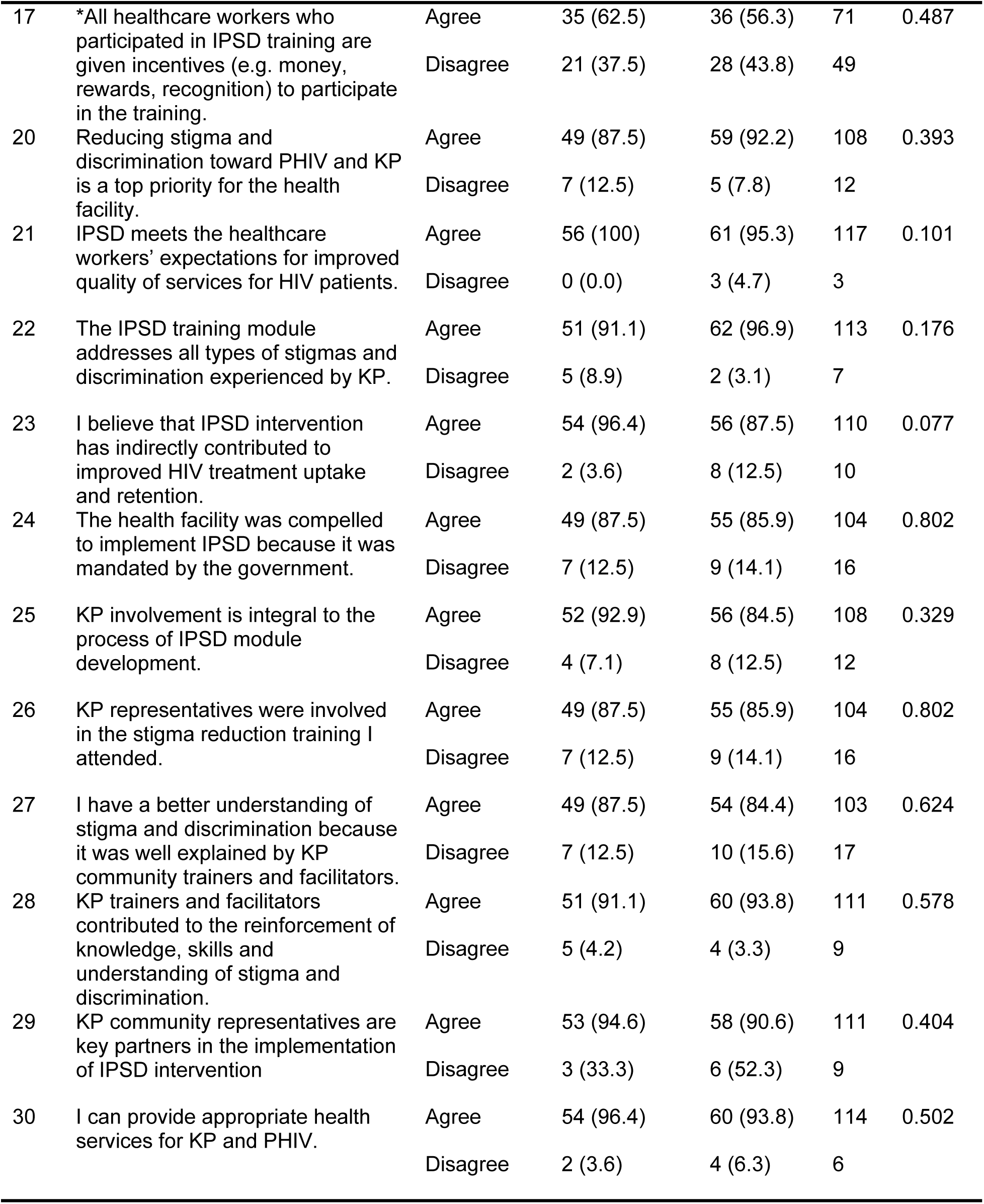
Association between Survey Items and PHCs Performance.

### Association of CFIR Constructs and Sub-constructs on Adoption

A comparison of the mean construct scores between HP- and AP-PHCs showed positive perceptions of the IPSD intervention across facilities, with no significant differences (*p* < 0.05; Table 3).

**Table 3:** Association between CFIR Domains, Constructs and Sub-Construct with PHC Performance (Adoption Outcome)

| Domain | Construct | Mean Score (SD) |  |  | *t* | *p*-value |
| --- | --- | --- | --- | --- | --- | --- |
|  |  | Overall<br>M(SD) | High<br>Performance<br>M(SD) | Average<br>Performance<br>M(SD) |  |  |
| **Inner Setting**<br>(number of items = 11) | **Tension for Change**<br>(number of items = 5) | 4.14 (0.50) | 4.20 (0.48) | 4.09 (0.52) | 1.157 | 0.25 |
|  | **Compatibility**<br>(number of items = 5) | 3.88 (0.53) | 3.90 (0.50) | 3.86 (0.56) | 0.412 | 0.681 |
|  | **Organisational Incentives and Rewards**<br>(number of items = 1) | 4.14 (0.50) | 3.80 (0.90) | 3.47 (0.98) | 1.942 | 0.055 |
| **Outer Setting**<br>(number of items = 5) | **Patient Needs and Resources**<br>(number of items = 4) | 4.16 (0.47) | 4.20 (0.50) | 4.12 (0.44) | 0.905 | 0.368 |
|  | **External Policy & Incentives**<br>(number of items = 1) | 4.02 (0.67) | 4.05 (0.72) | 3.98 (0.63) | 0.56 | 0.577 |
| **Process**<br>(number of items = 6) | **External Change Agents**<br>(number of items = 6) | 4.12 (0.46) | 4.18 (0.47) | 4.07 (0.44) | 1.27 | 0.207 |

Within the Inner Setting domain, HP-PHCs showed higher ratings for Tension for Change (M=4.20, SD=0.48) than AP-PHCs (M=4.09, SD=0.52; *p*=0.250). Compatibility was similar across groups. Organisational Incentives and Rewards showed the largest difference, with HP-PHCs (M=3.80, SD=0.90) compared to AP-PHCs (M=3.47, SD=0.98; *p*=0.055).

In the Outer Setting domain, both groups acknowledged Patient Needs and Resources, with HP-PHCs scoring higher (M=4.20, SD=0.50) than AP-PHCs (M=4.12, SD=0.44, *p*=0.368). External Policy and Incentives were consistently high across groups (*p*=0.577), reflecting a shared recognition of policy-level support.

Under the Process domain, HP-PHCs reported higher scores for External Change Agents (M=4.18, SD=0.47) than AP-PHCs (M=4.07, SD=0.44; *p*=0.207), suggesting stronger engagement with PWH and KP trainers. While no construct reached statistical significance, the findings suggest that perceptions related to organisational incentives, perceived need for change, and engagement with community actors may be important factors contributing to the adoption of IPSD.

### Fidelity Outcomes

#### Frequency Distribution Reported Against CFIR Constructs and Sub-Constructs

Overall fidelity to IPSD implementation was high, reflecting strong technical competence, supportive learning environments, and consistent post-training application (see Appendix 4).

#### Learning Climate

Most respondents (n=100, 83.2%) indicated adequate technical expertise within their PHCs to support IPSD implementation, and 73.4% (n=88) reported regularly engaging with clinical staff to improve practices. A supportive environment for discussing implementation challenges was evident (n=104, 86.6%), and ongoing collaboration with KPs (n=111, 92.5%) was reported.

#### Organisational Incentives and Rewards

Most (n=99, 82.5%) participants felt their efforts in stigma reduction were valued. Incentives were viewed as important for both initial adoption (n=73, 60.8%) and long-term adherence (n=76, 63.4%), underscoring the role of organisational support in sustaining fidelity.

#### Executing

Respondents demonstrated strong knowledge of Indonesia’s HIV/STI epidemiology (n=100, 83.3%) and national policies (n=104, 86.7%), with most (n=99, 82.5%) understanding the continuum of HIV care. Nearly all (n=112, 93.3%) credited KP-led training with improving their awareness of sexual orientation, gender identity, and expression (SOGIE). Awareness of stigma manifestations was also high (n=106, 88.4%), with most respondents recognising its harm to PWH and KP (n=112, 93.3%) and linking it to personal values and behaviours (n=98, 81.6%). Post-training, 84.2% (n=101) developed action plans, 75% (n=90) established monitoring systems, and 81.6% (n=98) pursued ongoing improvements in stigma reduction, indicating the active application of IPSD learnings.

#### Association between Survey Item Measuring Fidelity with PHCs Performance

Perceptions of intervention fidelity were generally consistent across facility types; however, significant differences were identified in two key indicators: respondents from HP-PHCs were more likely to report having adequate technical expertise to implement IPSD (n=51, 91.1%) compared with those from average-performing PHCs (n=49, 76.6%; *p*=0.033). Similarly, HP-PHC staff demonstrated stronger knowledge of HIV/AIDS and STI epidemiology (91.1% vs. 76.6%; *p*=0.033). Although the overall differences between the two groups were modest, the higher technical capacity and policy literacy in HP-PHCs likely contributed to more consistent and sustained fidelity in IPSD implementation (Table 4).

**Table 4:** Association between Survey Items Measuring Fidelity with PHCs Performance.

| Item | Statement | Agreed to the Statement, n (%) |  | p-value |
| --- | --- | --- | --- | --- |
|  |  | High Performance (N=56) | Average Performance (N=64) |  |
| 11 | We have enough technical expertise in our health facility to implement IPSD intervention. | 51 (91.1) | 49 (76.6) | 0.033 |
| 12 | There is regular engagement with clinical staff to discuss challenges and consider ways to improve the implementation of IPSD in our health facility. | 38 (67.9) | 50 (78.1) | 0.204 |
| 13 | The organisation provides a safe space (or a platform that protects our confidentiality) for us to address difficult issues and problems encountered in the implementation of IPSD. | 48 (85.7) | 56 (87.5) | 0.774 |
| 14 | The health facility is regularly engaged with KP representatives for consultation and support for the implementation of IPSD | 50 (89.3) | 61 (95.3) | 0.211 |
| 15 | All newly recruited healthcare workers are encouraged and supported to undertake IPSD training. | 52 (92.9) | 53 (82.8) | 0.097 |
| 16 | I am valued and receive recognition for my continued efforts in reducing HIV stigma and discrimination in the workplace. | 49 (87.5) | 50 (78.1) | 0.178 |
| 18 | Incentives (in any form) are necessary to motivate healthcare workers to adopt the IPSD intervention. | 37 (66.1) | 36 (56.3) | 0.272 |
| 19 | Incentives (in any form) are necessary to motivate healthcare workers to adhere to IPSD intervention. | 38 (67.9) | 38 (59.1) | 0.336 |
| 31 | I can explain the epidemic of HIV/AIDS and STIs in Indonesia. | 51 (91.1) | 49 (76.6) | 0.033 |
| 32 | I am aware of government policies relating to HIV/AIDS and STI control programmes in Indonesia. | 51 (91.1) | 53 (82.8) | 0.184 |
| 33 | I am aware of the laws and regulations related to the HIV/AIDS control program in Indonesia. | 49 (87.5) | 51 (79.7) | 0.252 |
| 34 | I can explain what the HIV Continuum of Care Strategy means. | 46 (82.1) | 53 (82.8) | 0.923 |
| 35 | I can identify the differences between gender, sex, and sexuality. | 53 (94.6) | 59 (92.2) | 0.591 |
| 36 | I can identify types and manifestations of stigma and discrimination towards KP and PWH in healthcare settings. | 51 (91.1) | 55 (85.9) | 0.382 |
| 37 | I understand the impact of stigma and discrimination toward KP and PHIV. | 52 (92.9) | 60 (93.8) | 0.845 |
|  |  | High Performance (N=56) | Average Performance (N=64) |  |
| 38 | I can explain the relationship between value systems, attitudes, and behaviour in expressing stigma and discrimination. | 44 (48.6) | 54 (84.4) | 0.412 |
| 39 | I can describe what a comprehensive package of health services for KP and PWH in health facilities means. | 45 (80.4) | 50 (78.1) | 0.764 |
| 40 | I am aware of potential stigmatising or discriminatory behaviours when providing health services to KP and PWH | 49 (87.5) | 56 (87.5) | 1.00 |
| 41 | After the IPSD training, the health facility developed an intervention plan to eliminate stigma and discrimination toward KP and PWH. | 45 (80.4) | 56 (87.5) | 0.285 |
| 42 | After the IPSD training, the health facility designed (or modified) a monitoring and evaluation system to monitor the delivery of the intervention. | 39 (69.6) | 51 (79.7) | 0.205 |
| 43 | My health facility develops continuous improvement efforts in the context of interventions to eliminate stigma and discrimination. | 46 (82.1) | 52 (81.3) | 0.900 |

#### Association of CFIR Constructs and Sub-Constructs on Implementation Fidelity

HCWs reported high fidelity in implementing the IPSD intervention across all CFIR constructs, with generally consistent perceptions between HP- and AP-PHCs. Execution received the highest overall rating (M=4.01, SD=0.48), with no significant difference between HP- and AP-PHCs (M=4.04 vs. 3.99; *p*=0.552). Learning Climate was also rated positively (M=3.97, SD=0.55), showing slightly higher but non-significant scores among HP-PHCs compared to AP-PHCs (M=4.03 vs. 3.93; *p*=0.368).

Organisational Incentives and Rewards were the only factors that demonstrated a statistically significant difference in PHC performance (*p*=0.032). Respondents from HP-PHCs reported stronger perceptions of institutional support and recognition (M=3.92, SD=0.68) than those from AP-PHCs (M=3.63, SD=0.81). This finding highlights the importance of organisational enablers, such as non-financial incentives, recognition, and leadership support, in enhancing implementation fidelity (Table 5).

**Table 5:** Association between CFIR Domains, Constructs and Sub-Constructs with PHC Performance (Fidelity Outcome)

| Domain | Construct | Mean Score (SD) |  |  | *t* | *p*-value |
| --- | --- | --- | --- | --- | --- | --- |
|  |  | Overall<br>M(SD) | High<br>Performance<br>M(SD) | Average<br>Performance<br>M(SD) |  |  |
| **Process**<br>(number of items=15) | **Executing**<br>(number of items=15) | 4.01 (0.48) | 4.04 (0.48) | 3.99 (0.48) | 0.596 | 0.552 |
| **Inner Setting**<br>(number of items=8) | **Learning Climate**<br>(number of items=5) | 3.97 (0.55) | 4.02 (0.49) | 3.93 (0.59) | 0.903 | 0.368 |
|  | **Organisational Incentives and Rewards**<br>(number of items=3) | 3.76 (0.76) | 3.92 (0.68) | 3.63 (0.81) | 2.169 | 0.032 |

## Discussion

These findings highlight the importance of reducing stigma in HIV care delivery through services that recognise, address, and mitigate stigma to ensure equitable, non-discriminatory, and person-centred services(11, 20). They align with multi-level stigma intervention evidence in LMICs showing that organisational climate and community engagement jointly enhance sustainability. High agreement across both HP- and AP-PHCs reflects consensus on the value of engaging PWH and KPs as co-implementers, consistent with evidence supporting contact-based strategies for knowledge translation in stigma reduction interventions (3, 10).

A significant association was found between occupational role and PHC performance (*p*=0.012). HP-PHCs had a higher proportion of doctors and nurses, whereas respondents from AP-PHCs were predominantly midwives and laboratory technicians. Although this pattern may reflect differences in staffing composition across facilities, it could also indicate sampling bias, with higher engagement from clinicians directly involved in IPSD implementation in HP-PHCs. Nonetheless, the presence of more clinically trained staff in these facilities may contribute to stronger leadership and a greater capacity to sustain intervention fidelity.

The IPSD intervention leveraged Tension for Change by creating an urgency for transformation, and its compatibility with existing workflows facilitated acceptance. These findings align with implementation science literature on organisational alignment in intervention delivery(21, 22).

Significant differences were observed in the Learning Climate and Executing constructs, both of which are core components of implementation fidelity. High-performing PHCs reported greater perceived technical capacity (*p*=0.033) and knowledge retention, particularly in HIV and STI epidemiology (*p*=0.033). These findings suggest that fidelity is influenced by a facility’s internal capacity to absorb and operationalise intervention components. While stigma-reduction content was delivered by KP and PHIV experts, the technical modules were facilitated by clinical professionals(16), which may have contributed to variations in how technical information was retained and applied across facilities.

The Organisational Incentives and Rewards sub-construct was used to assess both adoption and fidelity outcomes. While no significant difference was observed in adoption scores between HP- and AP-PHCs (*p*=0.25), fidelity scores differed significantly (*p*=0.032), with HP-PHCs reporting higher perceptions of organisational support and recognition. This finding suggests that beyond initial adoption, sustained implementation fidelity may be reinforced by effective capacity-building efforts and non-financial incentives that foster a more supportive and motivated work environment(23).

External policy influences were evident, as national mandates ensured uniform IPSD implementation across PHCs (*p*=0.802), underscoring the importance of government accountability and policy alignment in sustaining intervention fidelity(23, 24).

A more integrated view of the findings using the CFIR framework suggests that organisational and policy factors work together to influence implementation fidelity. Organisational incentives and a supportive learning climate (Inner Setting) help improve staff motivation and competence, while alignment with national policies and mandates (Outer Setting) strengthens accountability and consistency. These results indicate that maintaining fidelity depends not only on individual knowledge but also on the combined support from within the organisation and from external policies(23). Embedding CBPA principles within national HIV service accreditation processes could institutionalise stigma-reduction standards(25).

The study acknowledges several limitations. The challenges encountered in achieving the intended sample size may have compromised statistical power, and the geographic scope confined to selected districts within Greater Jakarta may limit the generalisability of the findings. As the data were self-reported, responses may have been subject to social desirability or recall bias, potentially leading to an overestimation of positive behaviours or attitudes. Additionally, the cross-sectional design restricted causal inference. Future research should adopt longitudinal designs to evaluate sustained impacts and conduct comparative studies across diverse settings to identify additional contextual factors that influence implementation success.

Given the limited sample, multivariable modelling was not feasible without compromising model validity and findings are exploratory. The achieved sample size (n=120) limits the study’s statistical power to detect small associations, and results should therefore be interpreted as indicative rather than confirmatory. Although the achieved sample was lower than the estimated 286, the study’s analytical focus remained descriptive and exploratory. Therefore, associations were explored descriptively within the CFIR framework to generate insights for future hypothesis testing. The findings are therefore interpreted with appropriate caution.

## Conclusion

This study provides insights into the IPSD intervention implementation, contributing to theory validation and extending implementation science frameworks like CFIR and Proctor et al. The findings indicate that organisational capacity, workforce composition, incentives, and community participation influence intervention adoption and fidelity. The results support CBPA’s effectiveness in enhancing stigma-reduction interventions within primary healthcare settings. This study demonstrates how participatory approaches improve intervention adoption and fidelity, particularly in resource-limited settings. Policymakers should prioritise strategies that strengthen these organisational and community factors for equitable HIV care. The study also shows CBPA’s adaptability across diverse contexts, establishing its potential for broader application in Southeast Asia and beyond.

## Data Availability

The datasets generated and/or analysed during the current study are not publicly available due to confidentiality agreements and ethical restrictions involving participant privacy. However, de-identified data may be made available from the corresponding author upon reasonable request and with approval from the relevant ethics committee. Any other identifying information related to the authors and/or their institutions, funders, approval committees, etc, that might compromise anonymity.

## Acknowledgements

We thank the staff at primary health centres across Greater Jakarta for their support in the data collection process and for their participation in the survey. Special appreciation goes to Dr. Rahmat Aji Pramono, HIV Program Coordinator at the Jakarta Provincial Health Office, as well as the District HIV Coordinators and Heads of Primary Health Centres, for their assistance in facilitating this research. This research was funded by the Malaysian Implementation Science Training program supported by Fogarty International Center, NIDA, NIMH and NICHHD (D43 TW-011324).

## Supporting Information

S1 Appendix 1: Selection of High and Average Performance Community Health Facilities

S2 Appendix 2: 5-Point Likert Scale Survey Instrument

S3 Appendix 3: Frequency Distribution (Adoption Outcome)

S4 Appendix 4: Frequency Distribution (Fidelity Outcome)

## Notes

### Competing Interest Statement

The authors have declared no competing interest.

### Funding Statement

Yes

### Author Declarations

This study was reviewed and approved by the Ethics Committee at the Institute of Research and Community Service, Atma Jaya Catholic University, Indonesia, under approval number 001D/III/PPE.PM.10.05/01/2023.

## References

1. UNAIDS. AIDSinfo UNAIDS Databook: Joint United Nations Programme on HIV/AIDS; 2024 [updated 2025. Available from: https://aidsinfo.unaids.org/databook.

2. UNAIDS. An evaluation of the contribution of the UNAIDS Joint Programme to strengthening HIV and primary health care outcomes: Indonesia. 2023. Report No.: UNAIDS/JC2996.

3. Link BG, Phelan JC. Conceptualizing Stigma. Annual Review of Sociology. 2001;27(1):363–85.

4. Fauk NK, Ward PR, Hawke K, Mwanri L. HIV Stigma and Discrimination: Perspectives and Personal Experiences of Healthcare Providers in Yogyakarta and Belu, Indonesia. Front Med (Lausanne). 2021;8:625787.

5. Rao D, Elshafei A, Nguyen M, Hatzenbuehler ML, Frey S, Go VF. A systematic review of multi-level stigma interventions: state of the science and future directions. BMC Med. 2019;17(1):41.

6. Fauk NK, Gesesew HA, Mwanri L, Hawke K, Ward PR. Understanding the quality of life of people living with HIV in rural and urban areas in Indonesia. PLoS One. 2023;18(7):e0280087.

7. Jaringan Indonesia Positif (JIP). People Living with HIV Stigma Index 2.0 in Indonesia. Jakarta: JIP; 2024.

8. FHI 360 FHI. HEALTH4ALL Training health workers for the provision of quality, stigma-free HIV services for key populationsLINKAGES. Available from: https://www.fhi360.org/resource/health4all.

9. IntraHealth International. The future of global health starts here: 7 creative approaches to health workforce challenges 2019 [Available from: https://www.intrahealth.org/sites/default/files/attachment-files/intrahealth7-creative-approaches-health-workforce-challenges.pdf.

10. Nickel S, von dem Knesebeck O. Effectiveness of Community-Based Health Promotion Interventions in Urban Areas: A Systematic Review. J Community Health. 2020;45(2):419–34.

11. Stangl A CD, Eckhaus T, Brady L, Nyblade L, Claeson M. Empowering communities to design and implement stigma-reduction programmes: Tackling HIV-related stigma and discrimination in South Asia 2010. Available from: https://documents1.worldbank.org/curated/en/463141468337182934/pdf/558410PUB0tack1C0disclosed071221101.pdf.

12. WHO. Health policy and system support to optimise community health worker programmes for HIV, TB and Malaria services: an evidence guide. World Health Organisation; 2020.

13. UNAIDS. Global AIDS Strategy 2021-2026, END INEQUALITIES. END AIDS.2021.

14. Proctor E, Silmere H, Raghavan R, Hovmand P, Aarons G, Bunger A, et al. Outcomes for implementation research: conceptual distinctions, measurement challenges, and research agenda. Adm Policy Ment Health. 2011;38(2):65–76.

15. Damschroder LJ, Reardon CM, Widerquist MAO, Lowery J. The updated Consolidated Framework for Implementation Research based on user feedback. Implement Sci. 2022;17(1):75.

16. Sheikh Mahmud M. The effectiveness of a community-based participatory approach (CBPA) in the implementation of an HIV stigma reduction intervention in Indonesia [Unpublished doctoral dissertation]. Kuala Lumpur, Malaysia: University of Malaya; 2025.

17. Indonesian AIDS Coalition (IAC). Community survey on access to “community-friendly HIV services” in Indonesia. [Unpublished report]. In press 2020.

18. Jaringan Indonesia Positif (JIP). Community satisfaction survey in primary health facilities [Unpublished report]. In press 2022.

19. Damschroder LJ, Aron DC, Keith RE, Kirsh SR, Alexander JA, Lowery JC. Fostering implementation of health services research findings into practice: a consolidated framework for advancing implementation science. Implement Sci. 2009;4:50.

20. Kemp CG, Jarrett BA, Kwon CS, Song L, Jette N, Sapag JC, et al. Implementation science and stigma reduction interventions in low- and middle-income countries: a systematic review. BMC Med. 2019;17(1):6.

21. Bertrand JT. Diffusion of innovations and HIV/AIDS. J Health Commun. 2004;9 Suppl 1:113–21.

22. Carlfjord S, Lindberg M, Bendtsen P, Nilsen P, Andersson A. Key factors influencing adoption of an innovation in primary health care: a qualitative study based on implementation theory. BMC Fam Pract. 2010;11:60.

23. Waluyo A, Culbert GJ, Levy J, Norr KF. Understanding HIV-related stigma among Indonesian nurses. J Assoc Nurses AIDS Care. 2015;26(1):69–80.

24. Edwards N, Kaseje D, Kahwa E, Klopper HC, Mill J, Webber J, et al. The impact of leadership hubs on the uptake of evidence-informed nursing practices and workplace policies for HIV care: a quasi-experimental study in Jamaica, Kenya, Uganda and South Africa. Implement Sci. 2016;11(1):110.

25. Murphy E, Shwe YY, Bhatia R, Bakkali T, Vannakit R. What will it take to end AIDS in Asia and the Pacific region by 2030? Sex Health. 2021;18(1):41–9.

